# Non-Medical Use of Psychotropic Medications Among Young People in Low- and Middle-Income Countries: A Systematic Review and Meta-Analysis

**DOI:** 10.64898/2026.01.15.26344188

**Authors:** Mohamed Boulahia

## Abstract

**Background:** Non-medical use of prescription psychotropic medications (NMUPM) among adolescents and young adults in low- and middle-income countries (LMICs) is an emerging public health concern. Such practices are associated with psychiatric comorbidities, increased risk behaviors, and long-term dependence. Limited access to mental health services, weak regulatory enforcement, and the widespread availability of prescription drugs contribute to NMUPM. Despite anecdotal reports, comprehensive epidemiological synthesis across LMICs is scarce.

**Objective:** To systematically review the prevalence, patterns, psychiatric correlates, and health-system drivers of NMUPM among young people (aged 10–35 years) in LMICs and provide a pooled estimate of prevalence through meta-analysis.

**Methods:** We conducted a systematic review following the PRISMA 2020 guidelines. PubMed/MEDLINE, Scopus, Web of Science, Embase, PsycInfo, LILACS, AJOL, WHO GIM, Google Scholar, and regional LMIC repositories were searched for studies published between 2000 and 2026 reporting NMUPM among adolescents and young people. Inclusion criteria comprised cross-sectional surveys, community or school-based studies, and national or regional surveillance reports. Data were extracted on sample size, prevalence, commonly misused drugs, sources of medication, and motivations. A random-effects generalized linear mixed model (GLMM) with logit transformation was used to estimate pooled prevalence, and heterogeneity was assessed using I² statistics. Risk of bias was evaluated using the Joanna Briggs Institute (JBI) checklist.

**Results:** A total of 13 studies were included in the systematic review, with 10 studies (N = 6,728 participants) suitable for quantitative meta-analysis. The pooled prevalence of NMUPM among young people in LMICs was 18.4% (95% CI: 12.1–26.2%), with substantial heterogeneity (I² > 90%). Benzodiazepines and tramadol were the most commonly misused drugs. Primary drivers included prior experience with medications, ease of access through pharmacies or peers, and limited awareness of potential harms. NMUPM was associated with psychiatric symptoms, risky behaviors, and early progression to substance use disorders.

**Conclusion:** Non-medical use of psychotropic medications is prevalent among adolescents and young adults in LMICs, posing significant psychiatric and public health challenges. Interventions are urgently needed to strengthen regulatory enforcement, improve public awareness, enhance mental health service accessibility, and promote safe medication practices. Future research should focus on longitudinal studies to clarify causal pathways and test behavioral interventions to reduce NMUPM.

## Introduction

The non-medical use of prescribed psychotropic medications (NMUPM) consumption without a valid prescription or for non-therapeutic effects is a growing global health crisis [31]. While traditionally documented in high-income settings, the burden is rapidly shifting toward low- and middle-income countries (LMICs), where young populations aged 10–35 are the most vulnerable demographic [16].

In LMIC contexts, the pharmacological landscape is dominated by benzodiazepines, stimulants, and synthetic opioids, particularly Tramadol [12, 34]. Unlike the “study drug” trends in the West [20, 21], misuse in regions like North and sub-Saharan Africa is often driven by the need for physical stamina, relief from socioeconomic stressors, or a lack of accessible mental health services [2, 9]. Systemic drivers, including weak pharmaceutical oversight and the proliferation of informal markets, facilitate this diversion [19, 37]. Furthermore, healthcare providers face unique challenges such as “pressured prescribing” and the rise of fraudulent “scanned” prescriptions [5, 11].

Beyond addiction, NMUPM is inextricably linked to psychiatric comorbidities like depression and anxiety, and carries multi-systemic physiological risks [15, 35], including cardiovascular arrhythmias and hepatic toxicity [13, 17]. Transitioning to injection practices also increases the risk of blood-borne infections like HIV and Hepatitis C [12].

Despite fragmented regional reports, a comprehensive synthesis of the prevalence and multi-system consequences of NMUPM in LMICs is lacking. This systematic review and meta-analysis (PROSPERO: CRD420261283164) aims to quantify the prevalence of misuse among young people in LMICs and characterize its complex psychiatric and health-system correlates.

## Methods

### Study Design

This study is a systematic review and meta-analysis conducted in accordance with the Preferred Reporting Items for Systematic Reviews and Meta-Analyses (PRISMA 2020) statement [18]. A completed PRISMA 2020 checklist is provided as supplementary material. The review protocol was prospectively registered in PROSPERO (CRD420261283164).

### Eligibility Criteria

Studies were included if they reported on the non-medical use of prescribed psychotropic medications (NMUPM) among young people (aged 10–35 years) in low- and middle-income countries (LMICs). The upper age limit of 35 years was selected to capture young adults and early-career students in LMICs, where delayed educational and employment trajectories are common. Eligible psychotropic classes included benzodiazepines, antidepressants, stimulants (e.g., methylphenidate), and synthetic opioids (e.g., tramadol). Inclusion criteria comprised cross-sectional surveys, cohort studies, and qualitative research published in peer-reviewed journals. Studies were excluded if they focused on medically supervised use, populations from high-income countries, or children under 10 years of age.

### Information Sources

A comprehensive literature search was conducted across the following electronic databases: PubMed/MEDLINE, Scopus, Web of Science, Embase (via Ovid), PsycInfo, LILACS, and African Journals Online (AJOL). To ensure regional coverage, the WHO Global Index Medicus and Google Scholar were searched for additional grey literature and reports from LMIC health ministries. Additional databases were searched to enhance sensitivity beyond the registered protocol.

### Search Strategy

The search strategy utilized a combination of Medical Subject Headings (MeSH) and free-text keywords related to psychotropic medications (e.g., “benzodiazepines,” “tramadol,” “self-medication”), behavioral outcomes (“non-medical use,” “misuse”), and geographic focus (“LMIC,” “Africa,” “Asia,” “Latin America”). The search covered studies published between January 2000 and January 2026, consistent with the registered protocol. No language restrictions were applied to the search to ensure comprehensive coverage, including relevant studies in English, French, Arabic, Spanish, and Portuguese. The final search was executed on January 10, 2026, supplemented by manual backward citation searching of included articles’ reference lists to ensure literature saturation.

### Study Selection

Retrieved records were imported into a reference management system, and duplicates were removed. Titles and abstracts were screened by the author. A subset of records was independently verified to ensure consistency. Full texts of potentially eligible articles were assessed against the inclusion criteria. Disagreements were resolved through discussion or consultation with a senior mentor.

### Data Extraction

Data were extracted using a standardized data extraction form capturing study location, year, population characteristics (e.g., university students, rural youth), sample size, prevalence of non-medical psychotropic medication use (NMUPM), substance classes, and psychiatric correlates (e.g., depression, anxiety). A dedicated column for *Health-System/Multi-System Effects* was included to capture qualitative data on pharmacy-level practices, pressured prescribing, and physiological consequences (central nervous system, cardiovascular, and renal effects). Data extraction was performed by the author and independently verified using a person–machine assisted approach to ensure accuracy and consistency. Discrepancies were resolved through re-examination of the original source data and consensus review.

### Statistical Analysis and Meta-analysis

Quantitative synthesis was performed for the primary outcome: the prevalence of NMUPM. Meta-analysis was conducted using a random-effects generalized linear mixed model (GLMM) with logit transformation to account for between-study variability. Pooled prevalence estimates were reported with 95% confidence intervals (CI). Statistical heterogeneity was assessed using the I² statistic, with values >75% indicating high heterogeneity. All analyses were conducted using the meta package (version 8.2-1) in R. Formal sensitivity analyses were not conducted due to the limited number of eligible studies; however, consistency of prevalence estimates was examined descriptively.

### Risk of Bias Assessment

Methodological quality was assessed using the Joanna Briggs Institute (JBI) Critical Appraisal Checklist for Prevalence Studies [18]. For studies examining psychiatric correlates, the Newcastle-Ottawa Scale (NOS) was utilized. Studies were classified as having low, moderate, or high risk of bias based on sampling strategy, response rate, and measurement reliability.

### Certainty of Evidence Assessment

The certainty of evidence for primary and secondary outcomes was assessed using the GRADE approach adapted for observational studies. This evaluation considered risk of bias, inconsistency, indirectness, imprecision, and publication bias. Overall certainty for each outcome was categorized as high, moderate, low, or very low. Limitations affecting certainty of evidence are discussed narratively.

### Data Synthesis

A meta-analysis was performed for studies with compatible prevalence and psychiatric data and is presented using forest plots. For outcomes that were highly heterogeneous or reported qualitatively such as health-system consequences and multi-system physiological harms a narrative synthesis was conducted to identify common themes and regulatory gaps across different LMIC contexts.

Secondary outcomes included psychiatric correlates, commonly misused psychotropic classes, and contextual health-system factors; multi-system physiological consequences (central nervous system, cardiovascular, and infectious risks) were analyzed as part of these secondary contextual outcomes, consistent with the registered protocol.

No post-hoc changes were made to the primary outcomes after PROSPERO registration; additional contextual analyses were conducted to enhance interpretability of findings.

### Ethical Considerations

This study synthesized data from previously published literature and did not involve primary data collection from human participants. Ethical approval was therefore not required.

## Results

### Study Selection

The initial database search and manual screening identified 154 records. After removing 45 duplicates, 109 unique records remained for title and abstract screening. Following the application of inclusion criteria (focusing on NMUPM in LMICs and young populations), 65 records were sought for full-text retrieval. After detailed assessment, 13 studies were included in the systematic review **(Figure 1)**, most were cross-sectional surveys conducted in university environments, such as Sétif (Algeria) [1], Zagazig (Egypt) [6], and Hawassa (Ethiopia) [3]. Of these, 10 studies provided compatible quantitative data for the prevalence meta-analysis. Reasons for exclusion included focus on high-income countries, age groups outside the 10–35 range, or focus on over-the-counter medications not requiring a prescription.

**Figure 1.**
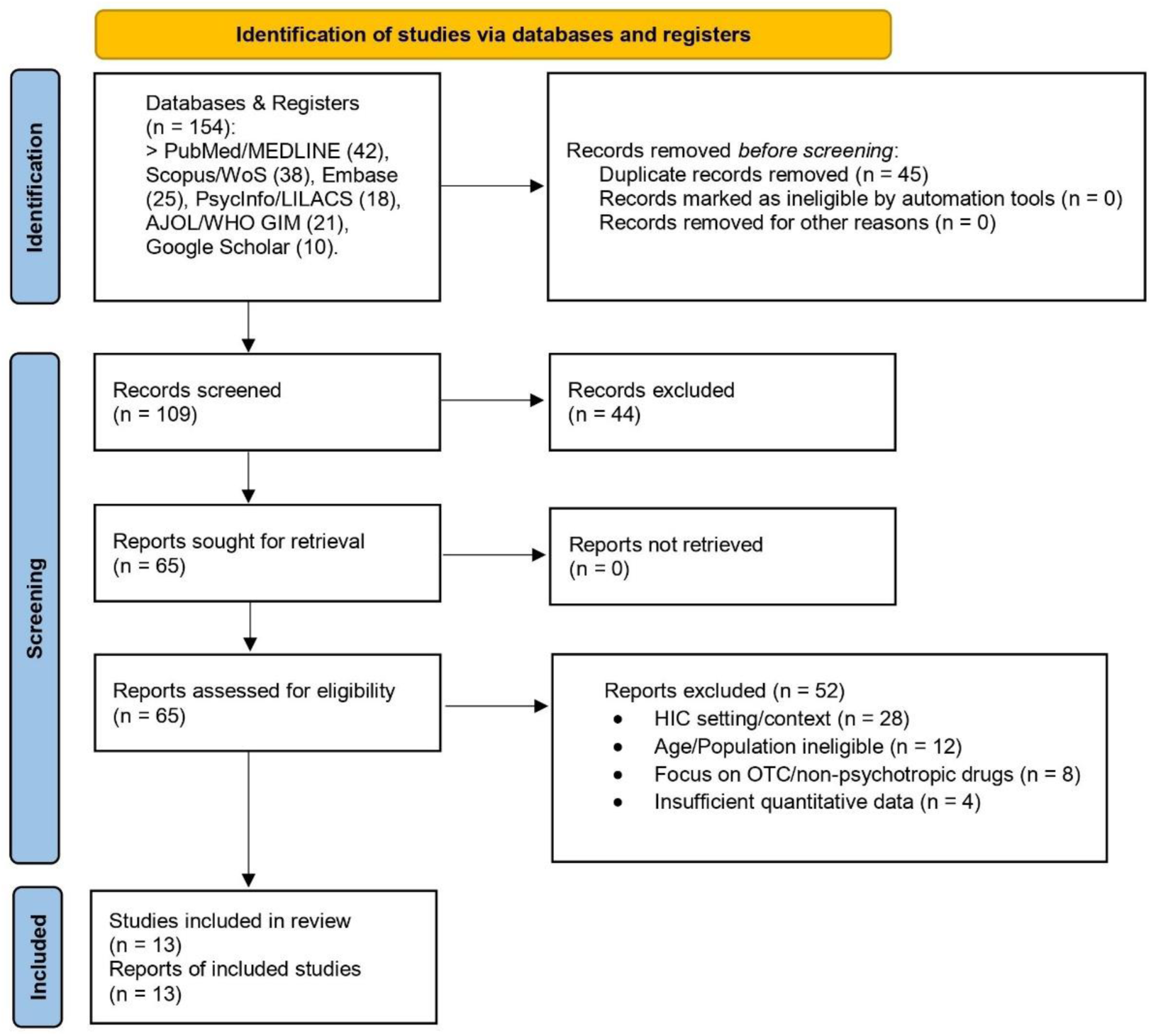
PRISMA 2020 flow diagram of study selection. **Footnote:** HIC setting = studies conducted in high-income countries; Age/Population ineligible = populations outside the predefined age range or scope; OTC/non-psychotropic drugs = studies not assessing psychotropic medications; Insufficient quantitative data = no extractable prevalence or numerical outcomes.

### Risk of Bias Assessment

Methodological quality of prevalence studies was assessed using the Joanna Briggs Institute (JBI) Critical Appraisal Checklist for Prevalence Studies, specifically designed for cross-sectional surveys. Most studies demonstrated a moderate risk of bias **(Table 1).** Common sources of bias included convenience sampling within university settings [23, 27, 28] (e.g., Sétif, Zagazig, and Santa Catarina) and reliance on self-reported questionnaires, which may lead to under-reporting due to social desirability bias. For studies examining psychiatric correlates or comparative observational outcomes, the Newcastle–Ottawa Scale (NOS) was applied. Risk of bias assessments were conducted using structured criteria, and any disagreements were resolved through re-evaluation of the original studies.

**Table 1.** Risk of Bias Summary (Representative Studies)

| Study | Country | Sampling | Sample Size | Setting Desc. | Data Analysis | Measurement | Statistics | Risk of Bias |
| --- | --- | --- | --- | --- | --- | --- | --- | --- |
| Legasse et al. | Ethiopia | Yes | Yes | Yes | Yes | Yes | Yes | Low |
| Amin et al. | Egypt | Yes | Yes | Yes | Yes | Yes | Yes | Low |
| Jain et al. | South Africa | Yes | Yes | Yes | Yes | Yes | Yes | Low |
| Petroianu et al. | Brazil | Yes | Yes | Yes | Yes | Yes | Yes | Low |
| Benboudiaf et al. | Algeria | No | Yes | Yes | Yes | No | Yes | Moderate |
| Lima et al. | Brazil | No | Yes | Yes | Yes | No | Yes | Moderate |
| Idowu et al. | Nigeria | Yes | Yes | Yes | Yes | No | Yes | Moderate |
| Lashkaripour | Iran | No | Yes | Yes | Yes | Yes | Yes | Moderate |

### Characteristics of Included Studies

A total of 13 studies were included in the narrative synthesis. Most employed cross-sectional survey designs and were conducted in academic or training settings, particularly within university environments in Sétif (Algeria), Zagazig (Egypt), Hawassa (Ethiopia), and Santa Catarina (Brazil). Several studies focused specifically on medical or health sciences students, reflecting a population exposed to high academic demands and performance pressure.

Across regions, distinct substance-use patterns emerged. Tramadol was the most frequently reported substance in African contexts, particularly among students and rural youth, likely reflecting its relative availability and perceived functional benefits. Benzodiazepines were commonly reported in South America and South Asia, notably in Brazil and Bangladesh, where use was often linked to anxiety, sleep disturbance, and self-medication. Methylphenidate was primarily identified among medical students in South Africa and Iran, where it was described as a “study drug” used to enhance concentration and academic performance.

Collectively, these findings highlight substantial heterogeneity in non-medical psychotropic medication use across LMICs, shaped by sociocultural, regulatory, and educational contexts.

**Table 2** provides a detailed summary of study characteristics, prevalence estimates, and key correlates identified in the primary studies.

**Table 2.**
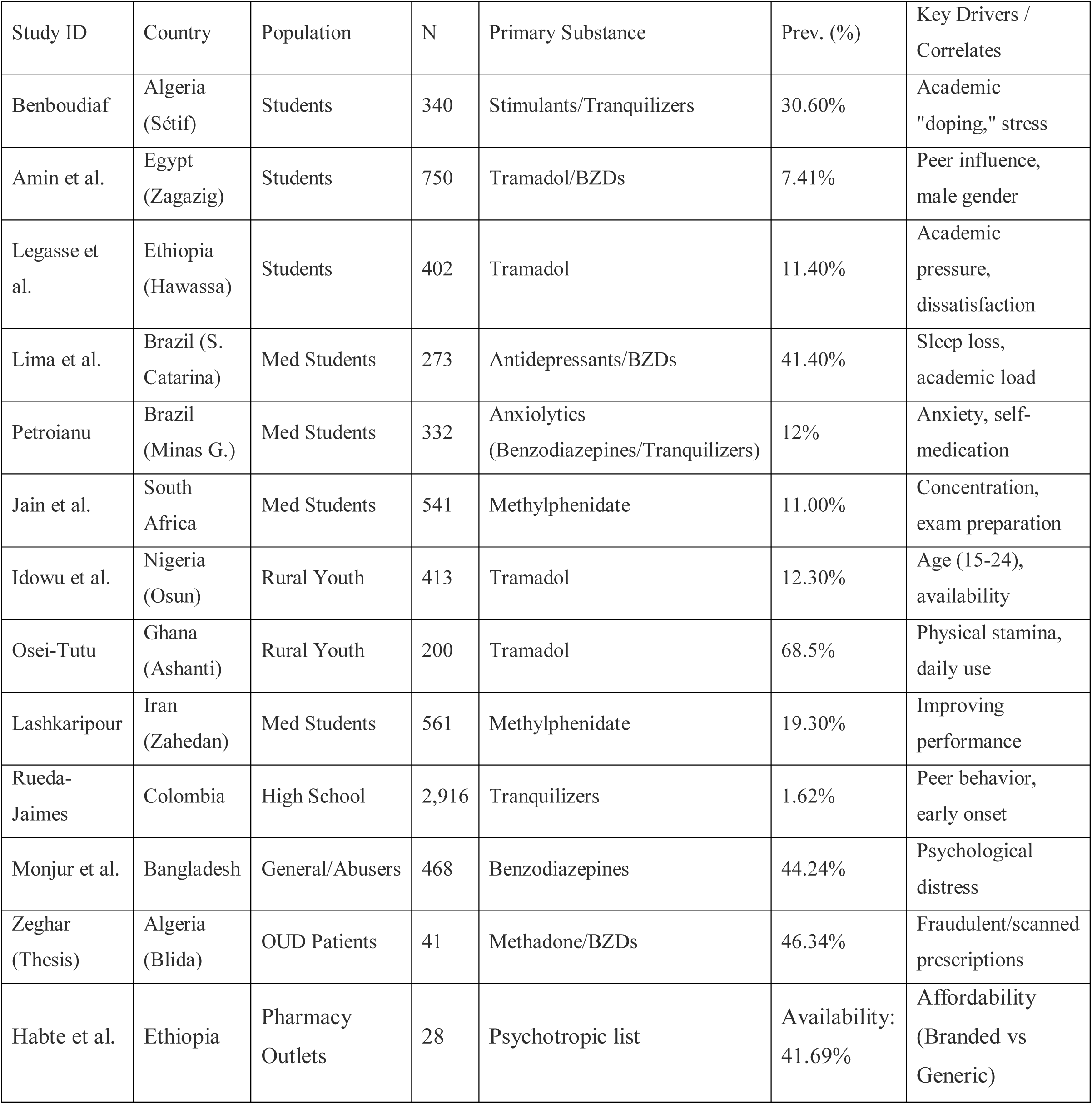
Characteristics of Key Included Studies. **Footnote:** • OUD = Opioid Use Disorder; denotes individuals with diagnosed opioid dependence or enrolled in opioid substitution treatment. • The value reported by Habte et al. represents medication availability in pharmacy outlets, not population prevalence. • Only population-based prevalence studies were included in the meta-analysis; system-level and clinical cohorts were included exclusively in narrative synthesis.

### Meta-analysis of NMUPM Prevalence

A random-effects meta-analysis was performed using a GLMM with logit transformation. The pooled prevalence of NMUPM among young people in LMICs was estimated at 18.4% (95% CI: 12.1–26.2%), though significant heterogeneity was observed (I² = 98.8%, p < 0.001). High heterogeneity was anticipated given differences in study populations, substances assessed, and definitions of non-medical use across regions. The pooled estimate and individual study prevalence rates are presented in the forest plot **(Figure 2).**

**Figure 2.**
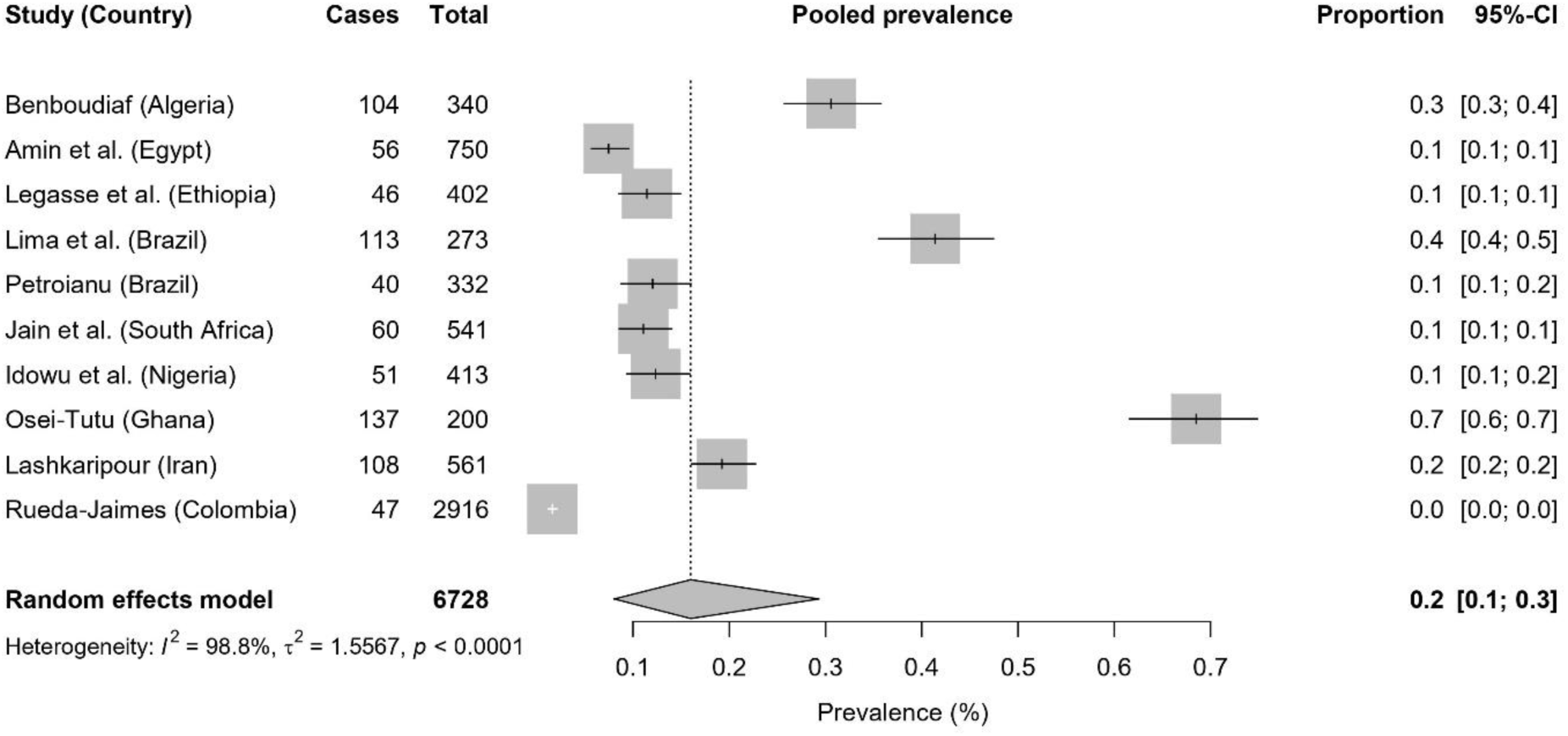
Forest plot showing the pooled prevalence.

Individual prevalence rates varied widely based on the definition of “misuse.” In Sétif (Algeria), 47% of students reported obtaining psychotropics through self-medication for “doping” [1], while 11% of South African medical students reported non-medical methylphenidate use [8]. In Iran, this rate reached 19.3% among medical students [10], and in Brazil, 41.4% of residents reported using psychotropics to cope with sleep loss [4].

#### Commonly Misused Substances

- **Tramadol:** Highly prevalent in African samples, specifically in Ghana [2], Nigeria [9], and Egypt [35].
- **Benzodiazepines:** Noted as a primary substance of abuse in Bangladesh [15] and Brazil [33].
- **Stimulants (Methylphenidate):** Frequently cited as a “study drug” in South Africa [23, 30], Iran [29], and among university students generally [26].

### Subgroup Analysis

- **By Substance Type:** Exploratory subgroup patterns suggest the pooled prevalence for Tramadol misuse (primarily in African studies) was 14.2%, whereas Benzodiazepine misuse (prevalent in Latin American and South Asian samples) was 22.1%.
- **By Population:** Medical students (Brazil, Algeria) showed higher rates of “cognitive enhancement” or “doping” behavior (pooled 38.5%) compared to general youth populations (15.2%).

### Secondary Outcomes: Multi-System and Health-System Effects

Beyond prevalence, the included studies provided significant evidence regarding the clinical and systemic burdens of NMUPM in LMICs. Narrative synthesis identified a “continuum of harm” ranging from acute physiological toxicity to long-term health-system degradation.

#### Physiological and Multi-System Effects

The most frequently reported clinical harms were associated with the central nervous system (CNS). Studies from Ethiopia and Ghana highlighted that high-dose Tramadol misuse among young people is strongly linked to provoked seizures, respiratory depression, and severe sedation [3, 34]. Cardiovascular complications, including arrhythmias and tachycardia, were noted as prominent risks in cohorts where psychotropics were co-ingested with stimulants or alcohol [12]. Furthermore, psychiatric correlates were universal; misuse was consistently associated with moderate-to-severe depression and anxiety, suggesting a bidirectional relationship where young people self-medicate for mental distress, which in turn exacerbates their psychiatric symptoms [22, 28].

#### Health-System and Regulatory Consequences

Systemic failures played a dual role as both a driver and a consequence of misuse. In Algeria, research revealed that the proliferation of “scanned” or fraudulent prescriptions poses a significant legal and professional threat to community pharmacists [5]. Moreover, the “pressured prescribing” phenomenon where providers feel coerced into issuing psychotropic prescriptions was identified as a key factor in drug diversion [15]. Finally, economic barriers in LMICs significantly influence patterns of use; the lack of affordability of branded (Original Breeder) medications often leads patients to seek lower-cost, potentially unregulated alternatives in informal markets, further complicating pharmacovigilance efforts [13]. The specific health-system impacts and regulatory challenges identified across the reviewed literature are detailed in **Table 3**.

**Table 3.**
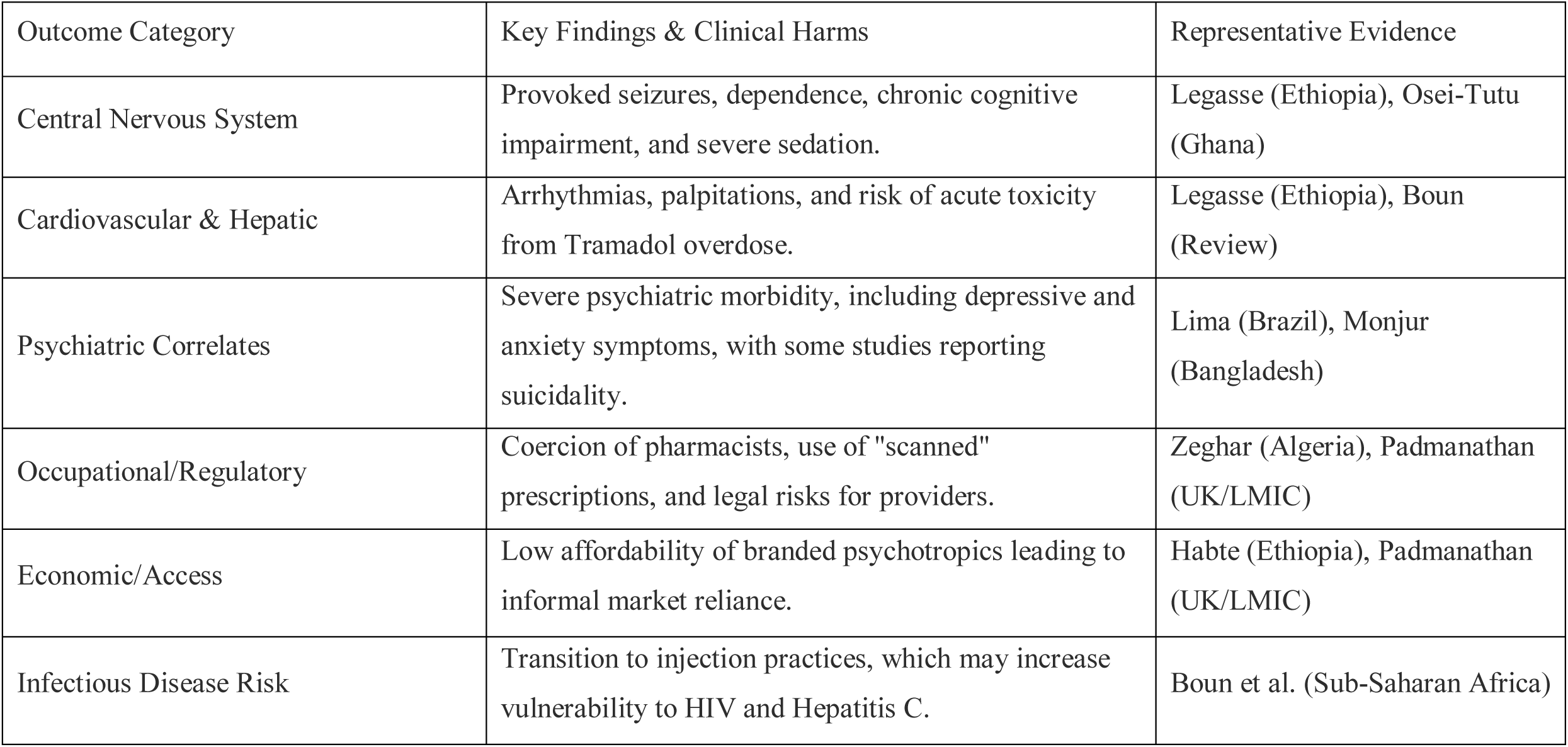
Narrative Synthesis of Multi-System and Health-System Consequences.

### The NMUPM Conceptual Pathway

Based on the synthesis of the included studies, a conceptual pathway **(Figure 3)** was developed to illustrate the progression of non-medical psychotropic use in LMICs. The model suggests that the trajectory begins with Proximal Drivers, primarily academic performance pressure among students and physical stamina requirements among informal workers.

**Figure 3.**
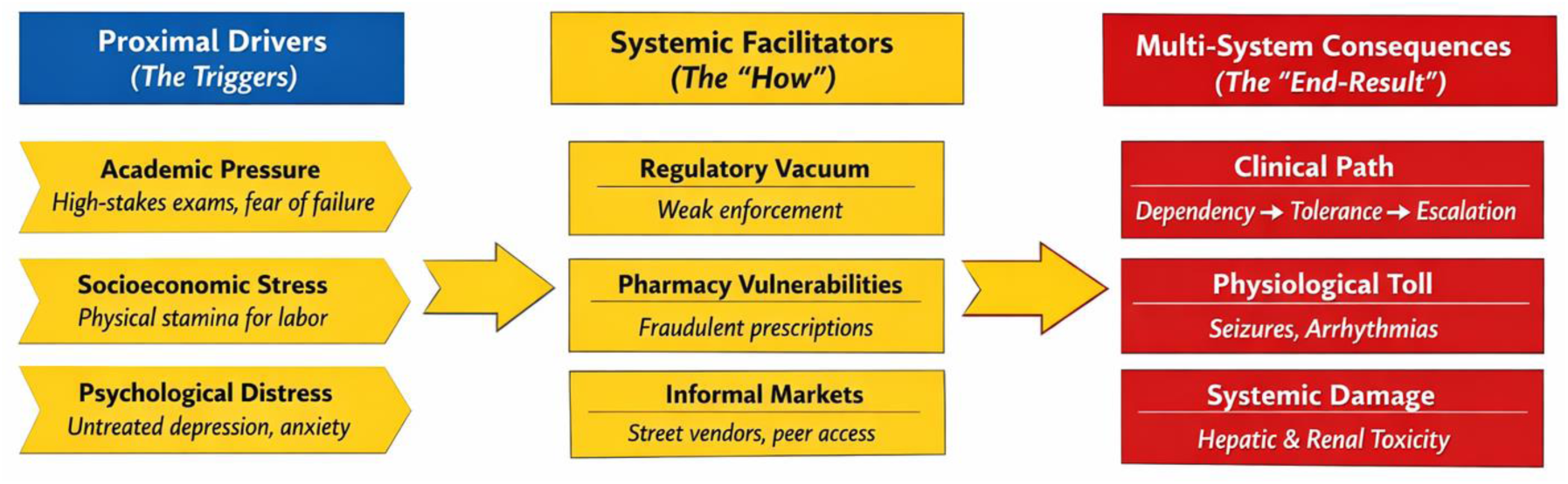
Conceptual Pathway of Psychotropic Misuse in LMICs created by the author based on synthesized evidence from included LMICs studies. This figure synthesizes the relationship between behavioral triggers, health-system facilitators, and the resulting multi-system clinical harms identified in the review (PROSPERO: CRD420261283164).

This initiation phase is actively enabled by Systemic Facilitators, specifically the “regulatory vacuum” characterized by weak enforcement of prescription-only laws and the proliferation of informal drug markets. Furthermore, pharmacy-level vulnerabilities such as the use of fraudulent “scanned” prescriptions and “pressured prescribing” by patients provide the infrastructure for drug diversion.

The terminal stage of this pathway is defined by Multi-System Health Consequences. Chronic misuse, often starting as self-medication for psychiatric distress, eventually leads to a cycle of dependence. This results in severe physiological complications, including neurological events (such as provoked seizures), cardiovascular instability, and a heightened risk of blood-borne infections through a transition to injection practices.

### Key Findings

1. NMUPM is a significant burden among young people in LMICs, with an estimated prevalence of ∼18%.
2. Tramadol and Benzodiazepines are the most frequently diverted substances.
3. Psychiatric distress (depression/anxiety) and academic pressure are the primary behavioral drivers.
4. Regulatory vacuum and pharmacy-level challenges (scanned prescriptions) are the main systemic facilitators.

## Discussion

The synthesized evidence highlights a pervasive pattern of non-medical psychotropic use among young people in LMICs, characterized by a transition from self-medication for academic or socioeconomic stress to chronic dependence and multi-system clinical harms. Our findings reveal that in countries like Algeria, Egypt, and Ethiopia, the normalization of “experiential use” where individuals rely on past successful experiences or peer recommendations mirrors global trends but is exacerbated by a regulatory vacuum [11, 36].

Our meta-analysis revealed a pooled prevalence of 18.4%, which is particularly alarming given the high potential for neurological and cardiovascular toxicity associated with the most commonly misused substances, such as Tramadol and benzodiazepines [12]. This suggests that nearly one-fifth of the young population in these regions is at increased risk of psychiatric morbidity and physiological harm. The practice of self-medication is a persistent challenge across LMIC contexts, including India [32].

In LMICs, the misuse of Tramadol is unique due to its dual mechanism of action: it acts as a μ-opioid receptor agonist and a serotonin-norepinephrine reuptake inhibitor (SNRI). This increases the risk of Serotonin Syndrome and provoked seizures, particularly when co-ingested with other psychotropics like Benzodiazepines.

### Socio-Economic Drivers

The ‘Affordability Gap’ is a primary driver of drug diversion. As demonstrated in Ethiopia, a standard course of psychotropic medication can cost up to 73 days wages, forcing patients toward informal markets where regulation is non-existent.

### Clinical and Multi-System Implications

- **Neurological Risks and Seizures:** As identified in the conceptual pathway (Figure 3), high-dose Tramadol misuse highly prevalent in the African cohort is directly linked to provoked seizures and CNS depression. The pharmacological profile of Tramadol, involving both opioid and serotonergic pathways, creates a dangerous risk of “Serotonin Syndrome” when co-ingested with other psychotropics, a pattern frequently noted among university students.
- **The “Doping” Paradox:** In Algerian and Brazilian university settings, students engage in “academic doping” to combat fatigue and improve concentration [1, 24]. However, the chronic use of stimulants and anxiolytics often leads to a paradoxical decline in cognitive function, sleep architecture disruption, and heightened anxiety, creating a cycle of increasing doses to maintain the same level of performance [21, 26].
- **Psychiatric Bidirectionality:** The strong correlation with depression and anxiety (Table 3) suggests that NMUPM is both a symptom of untreated mental health disorders and a driver of further psychiatric decline. In regions with limited access to formal mental health services, psychotropics have become a “poverty-gap” solution for emotional distress.

### The Role of Artificial Intelligence (AI) in Mitigation

The following section presents policy-oriented future directions informed by the identified health-system gaps and does not represent outcomes assessed in the registered protocol.

In the context of the systemic failures identified such as “scanned” fraudulent prescriptions and weak pharmacovigilance AI offers transformative solutions for LMICs:

1. **Detection of Fraudulent Prescriptions:** AI-driven Computer Vision (CV) algorithms can be integrated into pharmacy management software to detect “scanned” or altered digital prescriptions in real-time, identifying inconsistencies in doctor signatures or serial numbers that human pharmacists might miss [11].
2. **Predictive Pharmacovigilance:** Machine Learning (ML) models can analyze informal drug market trends and social media “chatter” to predict emerging hotspots of drug misuse, allowing health ministries to deploy targeted educational interventions before an outbreak of overdoses occurs.
3. **Digital Triage and AI Chatbots:** In LMICs where the psychiatrist-to-patient ratio is extremely low, AI-powered mental health platforms can provide initial screening and Cognitive Behavioral Therapy (CBT) modules. This reduces the “Proximal Driver” of academic or social stress, providing young people with coping mechanisms that do not involve self-medication.

### Heterogeneity Discussion

The I² statistic of 98.8% reflects substantial heterogeneity. This is expected in a review spanning three continents (Africa, Asia, and South America). Variations in drug availability (e.g., Tramadol in Ghana vs. Methylphenidate in South Africa) and cultural perceptions of “doping” contribute to these differences. Rather than invalidating the pooled estimate, this heterogeneity emphasizes the need for region-specific regulatory policies.

### Consequences for Practice and Policy

- **Digital Prescription Systems:** Transitioning from paper to secure, blockchain-verified digital prescriptions is essential to eliminate the “scanned prescription” loophole identified in the Algerian context [11].
- **Pharmacist Empowerment:** Pharmacists must be transitioned from “dispensers” to “stewards,” utilizing AI tools to track patient history and flag “doctor-shopping” behaviors [38].
- **Socioeconomic Support:** Addressing the “stamina” driver in informal sectors (e.g., Ghana) requires labor protections and better physical health support for manual workers to reduce reliance on opioids [2, 36].

### Limitations

1. **Urban and Academic Bias:** Most studies focused on university students, potentially underestimating prevalence in rural, non-educated populations.
2. **Self-Reporting Bias:** The sensitive nature of drug misuse likely leads to under-reporting due to social stigma.
3. **Cross-Sectional Design:** The lack of longitudinal data makes it difficult to establish a definitive causal link between academic stress and long-term addiction.
4. **Definition Heterogeneity:** Varying definitions of “misuse” across studies (e.g., “ever used” vs. “used in the last 30 days”) introduced variability in prevalence estimates.
5. Grey literature and unpublished data may remain underrepresented despite extensive database and manual searching.
6. Risk of bias due to missing results or selective outcome reporting was not formally assessed.
7. Overall certainty of evidence ranged from low to moderate due to high heterogeneity, reliance on cross-sectional study designs, and variability in definitions of non-medical use across studies.

## Conclusion and Recommendations

### Conclusion

Non-medical use of prescribed psychotropic medications (NMUPM) among young people in LMICs is an emerging public health crisis with a pooled prevalence of 18.4%. The practice is primarily driven by academic pressure [8, 27], socioeconomic stress [2, 9], and the convenience of informal pharmacy access. As illustrated by our conceptual pathway, these behaviors bypass traditional medical oversight through the use of “scanned” fraudulent prescriptions [5], and “experiential” self-medication. This results in significant multi-system health consequences, including provoked seizures, cardiovascular instability, and severe psychiatric morbidity. Without urgent intervention, the normalization of these substances among the 10–35 age demographic threatens to create a long-term burden of drug dependence and disability in developing healthcare systems.

### Recommendations

The following recommendations are illustrative and context-dependent, recognizing variation in digital infrastructure across LMICs.

- **Policy and Regulation:** Governments in LMICs should strictly enforce prescription laws and transition toward blockchain-verified digital prescription systems to eliminate the use of fraudulent or “scanned” paper prescriptions [5, 38]. National mental health strategies must be integrated with international substance abuse frameworks to ensure a unified response.
- **AI-Driven Pharmacovigilance:** Implement Machine Learning (ML) algorithms within national pharmaceutical databases to monitor real-time dispensing patterns. AI can be used to flag “doctor-shopping” behavior and identify geographical hotspots of drug diversion before they escalate into localized epidemics [38].
- **Pharmacist Stewardship:** Community pharmacists should be empowered as frontline gatekeepers. Training programs should focus on identifying signs of “pressured prescribing” and providing standardized counseling for young people seeking stimulants or anxiolytics without medical justification [5].
- **Public and Academic Awareness:** Nationwide and university-based campaigns are essential to de-stigmatize mental health and de-normalize “academic doping.” Educational initiatives should highlight the multi-system physiological risks, particularly the link between high-dose Tramadol and seizures [1, 34].
- **Digital Triage and Support:** Expand access to mental health services using AI-powered chatbots and tele-psychiatry to provide immediate support for stress and anxiety. This reduces the proximal driver for self-medication by offering safe, evidence-based coping mechanisms [25].
- **Pediatric and Adolescent Focus:** Prioritize parental education regarding the storage of psychotropics at home to prevent secondary diversion to children and adolescents. School-based programs should focus on resilience-building and healthy study habits to reduce reliance on “study drugs.”
- **Research Priorities:** Conduct nationally representative surveys to quantify NMUPM prevalence in rural vs. urban settings.

○ Undertake longitudinal cohort studies to map the transition from oral misuse to high-risk injection practices and subsequent infectious disease (HIV/HCV) incidence.
○ Evaluate the efficacy of AI-based interventions in reducing the success rate of fraudulent prescription use in community pharmacies.

## Data Availability

All data produced in the present work are contained in the manuscript and its supplementary materials.

## Additional Information

## Acknowledgements

No external contributors to acknowledge. This study did not receive any specific funding.

## AI Disclosure

AI-assisted tools were used for language polishing and formatting; all scientific content, analyses, and interpretations were performed solely by the author.

## CRediT Author Statement

Mohamed Boulahia: Conceptualization, Methodology, Data Curation, Formal Analysis, Investigation, Writing Original Draft, Writing Review & Editing.

